# Efficacy and safety of bempedoic acid in women with hypercholesterolemia: Pooled analyses from phase 3 trials

**DOI:** 10.1101/2023.02.14.23285912

**Authors:** Anne C. Goldberg, Maciej Banach, Alberico L. Catapano, P. Barton Duell, Lawrence A. Leiter, Jeffrey C. Hanselman, Lei Lei, G.B. John Mancini

## Abstract

**Background and aims:** Sex-specific differences in the response to lipid-lowering therapies have been reported. Here, we assessed the effect of bempedoic acid in women and men using pooled, patient-level data from four phase 3 clinical trials of bempedoic acid.

**Methods:** Patients were grouped into two pools: atherosclerotic cardiovascular disease (ASCVD) and/or heterozygous familial hypercholesterolemia (HeFH) “on statins”, and “low-dose or no statin”. Percent changes from baseline to at least week 12 in low-density lipoprotein-cholesterol (LDL-C), non–high-density lipoprotein-cholesterol (non–HDL-C), total cholesterol (TC), apolipoprotein B (Apo B), and high-sensitivity C-reactive protein (hsCRP), as well as safety, were analyzed by statin pool and sex.

**Results:** Overall, 3623 patients were included (bempedoic acid, 2425; placebo, 1198). Significant reductions in lipid parameters and hsCRP were observed with bempedoic acid *vs*. placebo in both sexes in the ASCVD and/or HeFH on statins (n = 3009) and the low-dose or no statin (n = 614) pools (*p*≤0.002). Compared with men, women had significantly greater placebo-corrected reductions in LDL-C (−21.2% *vs*. −17.4%; *p*=0.044), non–HDL-C (−17.3% *vs*. −12.1%; *p*=0.003), TC (−13.8% *vs*. −10.5%; *p*=0.012), and Apo B (*−*16.0% *vs*. −11.3%; *p*=0.004) in the ASCVD and/or HeFH on statins pool. Women had numerically greater reductions than men in lipid parameters in the low-dose or no statin pool and hsCRP in both pools. The safety of bempedoic acid was comparable between sexes.

**Conclusions:** In this pooled analysis, women experienced significant improvements in levels of LDL-C and other lipid parameters with bempedoic acid.

## Introduction

Cardiovascular disease (CVD) contributes to a considerable disability and mortality burden and is the leading cause of death in the US and worldwide [1, 2]. While women and men have a comparable overall lifetime risk of CVD [3], there are significant sex-related differences in the manifestation of the disease. Age-adjusted CVD mortality is higher in men than women in the US [4]. However, women have higher mortality and worse prognosis than men after acute cardiovascular events [5].

Women are generally less likely to benefit from healthcare utilization than men in the primary and secondary prevention of CVD, which may be due to underdiagnosis, underuse of screening, and the misconception that CVD is a problem that primarily affects men [1]. Women are less likely than men to receive guideline-recommended lipid-lowering therapies (LLTs), such as statins, and consequently less likely to achieve recommended low-density lipoprotein cholesterol (LDL-C) levels [6-9]. Women and men with a comparable risk of major vascular events derive similar cardioprotective benefits from the same level of LDL-C lowering with LLTs [10]. Despite this, women are more likely to discontinue using, or to not use, statin therapy [9, 11, 12], which may be related in part to women being more likely than men to experience statin-associated adverse events, primarily involving muscle symptoms [12, 13]. Non-statin LLTs may therefore be more frequently required for women to achieve LDL-C recommended levels. Additionally, women with primary hypercholesterolemia and mixed dyslipidemia who are treated with niacin, experience greater changes in lipids than men [14]. Considering these observations, sex-specific responses should be evaluated for other LLTs.

Bempedoic acid is a first-in-class oral inhibitor of ATP citrate lyase (ACL), an enzyme upstream of 3-hydroxy-3-methylglutaryl-coenzyme A (HMG-CoA)—the target of statins—in the cholesterol biosynthetic pathway [15]. Bempedoic acid is a prodrug that is activated in the liver by very long-chain acyl-CoA synthetase-1 (ACSVL1), leading to the reduction of cholesterol synthesis and upregulation of hepatic low-density lipoprotein (LDL) receptors that mediate increased uptake and clearance of plasma LDL [15].

The efficacy of bempedoic acid in lowering LDL-C has been demonstrated in phase 3 clinical trials [16-19]. Analyses of pooled data from four phase 3 trials of bempedoic acid *vs*. placebo demonstrated a possible difference in observed reductions of LDL-C between women and men [20, 21]. Women tended to have greater reductions in LDL-C than men both in patients with atherosclerotic cardiovascular disease (ASCVD) or heterozygous familial hypercholesterolemia (HeFH) (−21.2% *vs*. −17.4%, *p*=0.04) and in patients with hypercholesterolemia and statin intolerance (−27.7% *vs*. −22.1%, *p*=0.08) [20]. Moreover, a separate post-hoc analysis showed that among patients with high cardiovascular risk who received bempedoic acid, women were 1.6 times more likely than men to achieve a minimum of 30% reduction in LDL-C at week 12 from baseline (*p*<0.0001) [21]. Here, we further evaluate and compare sex-specific efficacy and safety of bempedoic acid in women and men who required additional LDL-C lowering, using patient-level pooled data from the same four phase 3, placebo-controlled randomized clinical trials of bempedoic acid.

## Patients and methods

### Study design

This study is a post-hoc analysis of patient-level, pooled data from four phase 3 clinical trials that evaluated the safety and efficacy of bempedoic acid. Each of the four trials has been previously described [16-19]. CLEAR Wisdom (NCT02991118) [17], CLEAR Harmony (NCT02666664) [16], CLEAR Tranquility (NCT03001076) [18], and CLEAR Serenity (NCT02988115) [19] were randomized, double-blind, placebo-controlled, parallel-group, multicenter, phase 3 studies. In all four studies, adult patients with hypercholesterolemia on stable LLT requiring additional LDL-C lowering were randomized 2:1 to bempedoic acid 180 mg once daily or placebo. Women enrolled were either naturally postmenopausal, surgically sterile, or of childbearing potential and willing to use two acceptable methods of birth control.

The CLEAR Harmony and CLEAR Wisdom studies were both 52-week studies that enrolled patients with established ASCVD and/or HeFH, receiving stable maximally tolerated doses of statins, with or without other approved LLT [16, 17]. Maximally tolerated statin was defined as the highest intensity statin regimen that a patient could maintain, including low-dose or no statin, but excluding simvastatin at daily doses of 40 mg or greater, and was determined by the principal investigator (Supplementary Table 1) [16, 17]. Statin intensity classification was based on the 2013 American College of Cardiology/American Heart Association guidelines [16, 17, 22].

The 24-week CLEAR Serenity and the 12-week CLEAR Tranquility studies enrolled patients with a history of statin intolerance who were taking no greater than low-dose statins [18, 19]. Low-dose statin therapy was defined as an average daily dose of rosuvastatin 5 mg, atorvastatin 10 mg, simvastatin 10 mg, lovastatin 20 mg, pravastatin 40 mg, fluvastatin 40 mg, or pitavastatin 2 mg; doses lower than these were considered very low–dose statin therapy (Supplementary Table 1) [18, 19].

All studies were conducted in accordance with the Declaration of Helsinki and Good Clinical Practice guidelines. All protocols were approved by local independent ethics committees at each study site, and all study participants provided written informed consent.

### Endpoints and assessments

Efficacy and safety endpoints of the four phase 3 trials have been previously described [16-19]. All four studies assessed the percent change from baseline to at least week 12 in levels of LDL-C, non–high-density lipoprotein-cholesterol (non–HDL-C), total cholesterol (TC), apolipoprotein B (Apo B), and high-sensitivity C-reactive protein (hsCRP) as prospective efficacy endpoints. Clinical laboratory samples were collected up to week 52 in CLEAR Wisdom and CLEAR Harmony, and up to week 24 and week 12 for CLEAR Serenity and CLEAR Tranquility, respectively [16-19]. Quantification of plasma lipid levels and other parameters has been previously described [20]. Safety evaluations included the occurrence of treatment-emergent adverse events (TEAEs) up to 30 days after the end of treatment.

### Statistical analysis

The efficacy analysis population included all patients who were randomized to treatment; the safety analysis population included all patients receiving at least one dose of study treatment. To control for differences in patient characteristics and background therapy, patients were grouped into two pools based on the similarity of the study designs: 1) the ASCVD and/or HeFH “on statins” pool, which included patients from CLEAR Harmony and CLEAR Wisdom, and 2) the “low-dose or no statin” pool, which included patients from CLEAR Tranquility and CLEAR Serenity. All endpoints were analyzed by statin pool and female or male sex assigned at birth. No imputation was performed for missing data. Treatment effects were presented as least squares (LS) means, estimated by an analysis of covariance (ANCOVA) model, with percent change from baseline as the dependent variable, study and treatment as fixed factors, and baseline as a covariate. Comparisons of hsCRP were based on a nonparametric method. The interaction *p* value was obtained from the interaction term via an ANCOVA model with percent change from baseline as the dependent variable (log-transformed for hsCRP), study, treatment, sex, and interaction term between treatment and sex as fixed factors, and baseline as a covariate, using α = 0.05. No multiplicity adjustment was performed, and *p* values presented are nominal. For the CLEAR Harmony and CLEAR Wisdom trials, mean levels of LDL-C, non– HDL-C, TC, Apo B, and hsCRP were plotted from baseline up to week 52. Adverse events were coded per the Medical Dictionary for Regulatory Activities (MedDRA) version 20.1 and summarized as patient incidences.

## Results

### Patients

Overall, 3623 and 3621 patients were included in the pooled efficacy and safety analyses, respectively. Within the pooled efficacy population, 2425 patients were treated with bempedoic acid and 1198 received placebo. There were 3009 patients in the ASCVD and/or HeFH on statins pool and 614 patients in the low-dose or no statin pool. In the ASCVD and/or HeFH on statins pool, 91.1% of patients were on a moderate or high-intensity statin. Across both pools, 1244 (34.3%) patients were women and 2379 (65.7%) were men. There were more women than men in the low-dose or no statin pool (58.5%) and fewer women than men in the ASCVD and/or HeFH on statins pool (29.4%).

Baseline demographics were generally comparable between women and men across both treatment groups and statin pools (Supplementary Table 2). Black race and Hispanic ethnicity were more prevalent among women than men in the ASCVD and/or HeFH on statins pool, although the absolute numbers were small. There were more women than men with HeFH in the ASCVD and/or HeFH on statins pool across both treatment groups. Across treatment groups and statin pools, ≤5.2% of women were premenopausal. Among non-premenopausal women, ≤10% were taking hormone replacement therapy (HRT) across both treatment groups and statin pools. Hypertension was less prevalent among women than men in the low-dose or no statin pool. Overall, baseline lipid and hsCRP concentrations were slightly higher in women *vs*. men across both treatment groups in the ASCVD and/or HeFH on statins and the low-dose or no statin pools, apart from triglycerides in the low-dose or no statin pool.

### Efficacy endpoints

Significant reductions from baseline in LDL-C at week 12 were observed for patients who received bempedoic acid compared with those who received placebo (*p*<0.001, **Fig. 1**). In both pools, women had greater placebo-corrected reductions in LDL-C than men. The difference in the magnitude of LDL-C lowering between women and men was statistically significant in the ASCVD and/or HeFH on statins pool (−21.2% *vs*. −17.4% for women *vs*. men, interaction *p*=0.044) but not in the low-dose or no statin pool (−27.7% *vs*. −22.1%, for women *vs*. men, interaction *p*=0.079; **Fig. 1**). Among patients in the low-dose or no statin pool who were not on statins, women had significantly greater placebo-corrected reductions in LDL-C than men (−30.5% *vs*. −22.6% for women *vs*. men; interaction *p*=0.018); LDL-C reductions among patients on low-dose statins were similar between women and men (Supplementary Figure 1). Among patients who received bempedoic acid, the majority of women and men in the ASCVD and/or HeFH on statins pool achieved LDL-C levels lower than 100 mg/dL (68.7% and 76.2%, respectively). Conversely, fewer than half of women and men in the low-dose or no statin pool achieved this threshold (46.2% and 42.4%, respectively; **Fig. 2**).

**Fig. 1.**
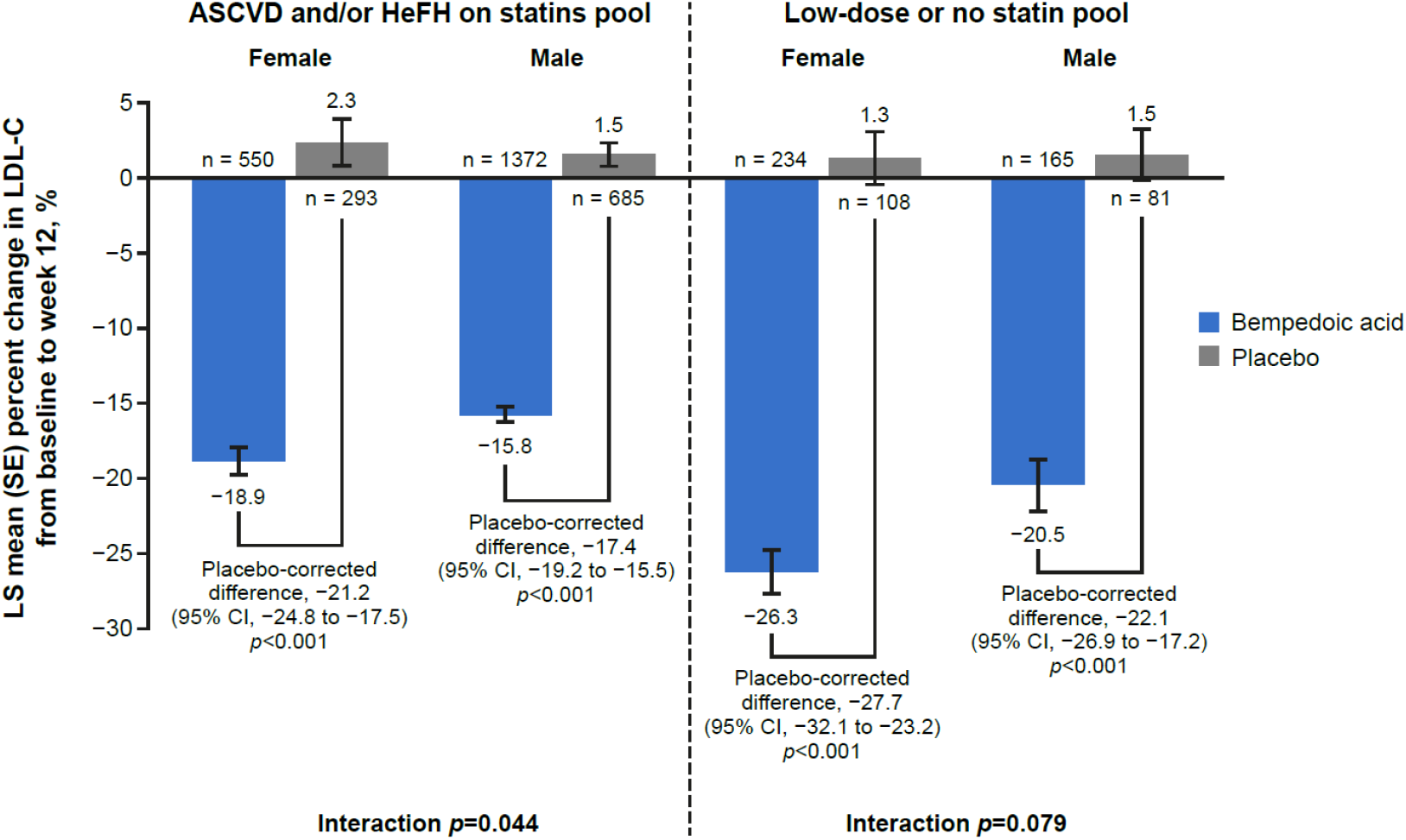
Percent change from baseline to week 12 and placebo-corrected difference (95% CI) in LDL-C by statin pool and sex at birth. ASCVD, atherosclerotic cardiovascular disease; CI, confidence interval; HeFH, heterozygous familial hypercholesterolemia; LDL-C, low-density lipoprotein cholesterol; LS, least squares; SE, standard error. Numbers on the x axis represent the total number of patients within each group with available LDL-C data at week 12.

**Fig. 2.**
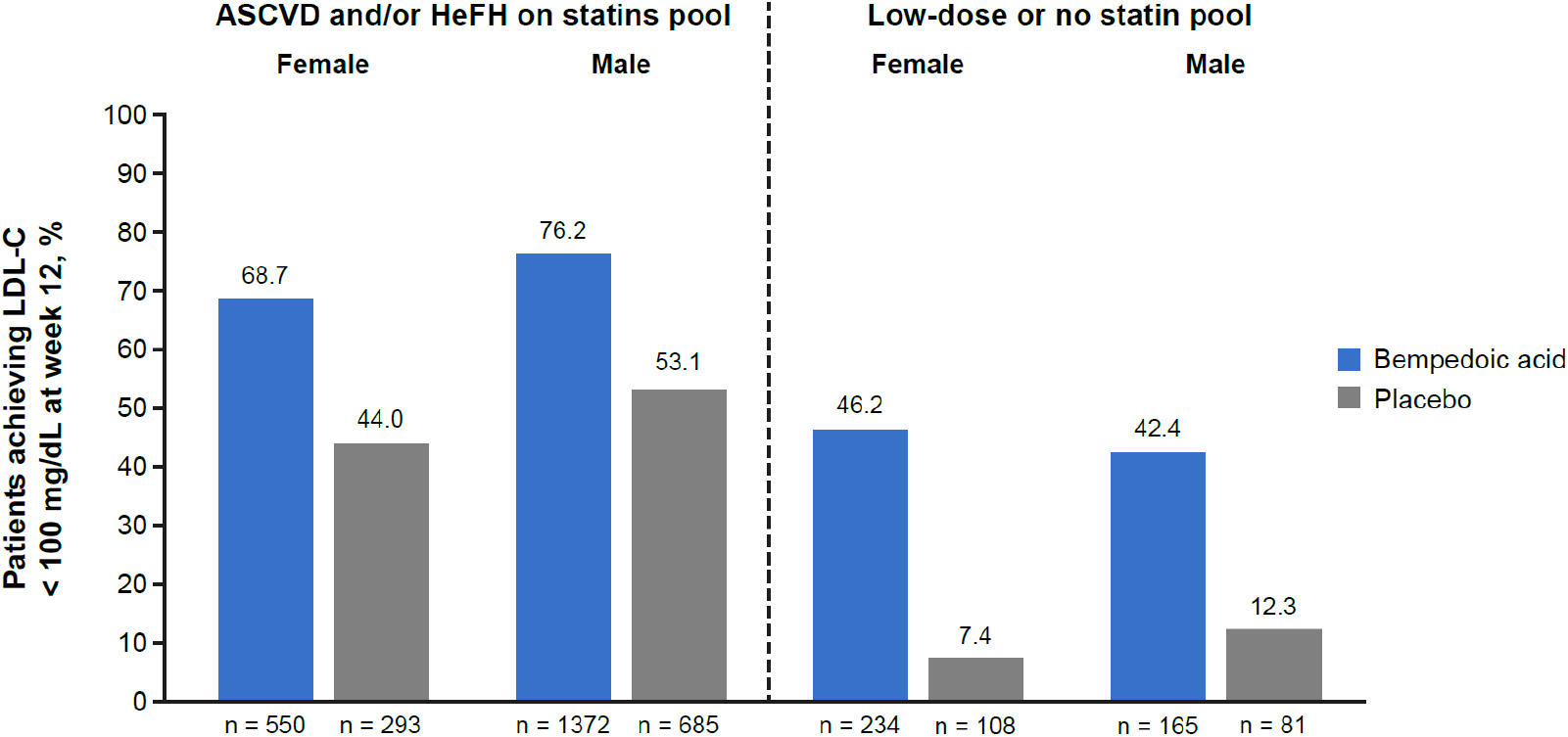
Proportion of patients achieving LDL-C levels <100 mg/dL at week 12 by statin pool and sex at birth. Numbers on the x axis represent the total number of patients within each group with available LDL-C goal attainment data at week 12. ASCVD, atherosclerotic cardiovascular disease; HeFH, heterozygous familial hypercholesterolemia; LDL-C, low-density lipoprotein cholesterol.

Significant reductions were also observed in TC, non–HDL-C, Apo B, and hsCRP levels from baseline to week 12 in patients receiving bempedoic acid compared with placebo in both sexes from both statin pools (*p*≤0.002; **Fig. 3**). Women experienced significantly greater placebo-corrected reductions in TC (−13.8% *vs*. −10.5%, interaction *p*=0.012), non–HDL-C (−17.3% *vs*. −12.1%, interaction *p*=0.003), and Apo B (−16.0% *vs*. −11.3%, interaction *p*=0.004) than men and numerically greater reductions in hsCRP (−20.0% *vs*. −17.5%, interaction *p*>0.05) in the ASCVD and/or HeFH on statins pool. In the low-dose or no statin pool, women experienced numerically greater placebo-corrected reductions in all lipid parameters and hsCRP than men (interaction *p*>0.05).

**Fig. 3.**
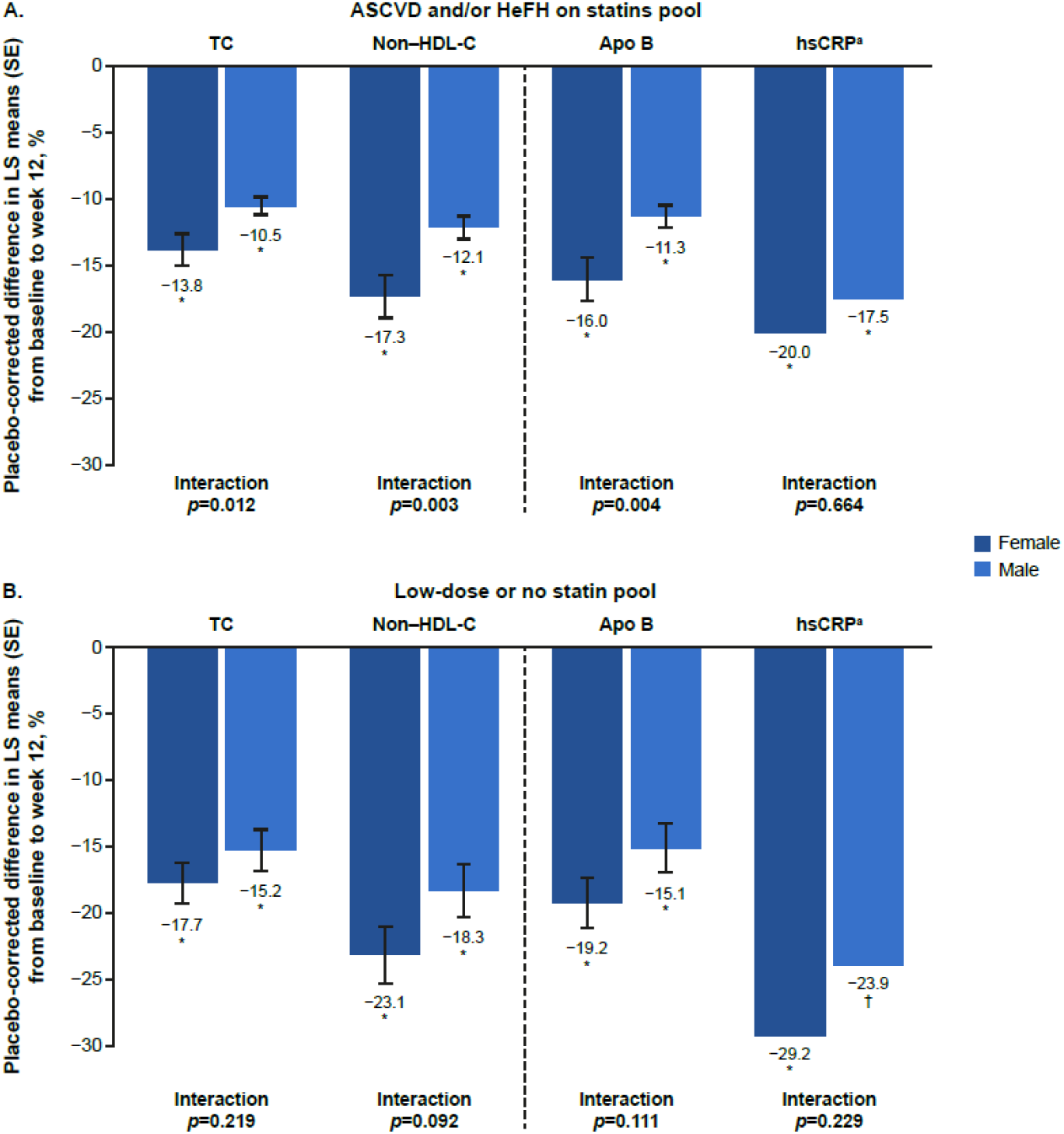
Percent changes in lipid parameters and hsCRP by statin pool and sex at birth. Placebo-corrected percent changes in TC, non–HDL-C, Apo B, and hsCRP from baseline to week 12 by sex in the (A) ASCVD and/or HeFH on statins pool and (B) low-dose or no statin pool. Apo B, apolipoprotein B; ASCVD, atherosclerotic cardiovascular disease; HeFH, heterozygous familial hypercholesterolemia; hsCRP, high-sensitivity C-reactive protein; LS, least squares; non–HDL-C, non–high-density lipoprotein cholesterol; SE, standard error; TC, total cholesterol. \**p*<0.001 for comparisons *vs*. placebo. ^†^ *p*=0.002 for comparisons *vs*. placebo. aData for hsCRP are presented as location shift.

In the ASCVD and/or HeFH on statins pool, the effect of bempedoic acid on LDL-C, non– HDL-C, TC, Apo B, and hsCRP was still apparent through week 52, with minimal attenuation of effect over time regardless of sex (**Fig. 4**).

**Fig. 4.**
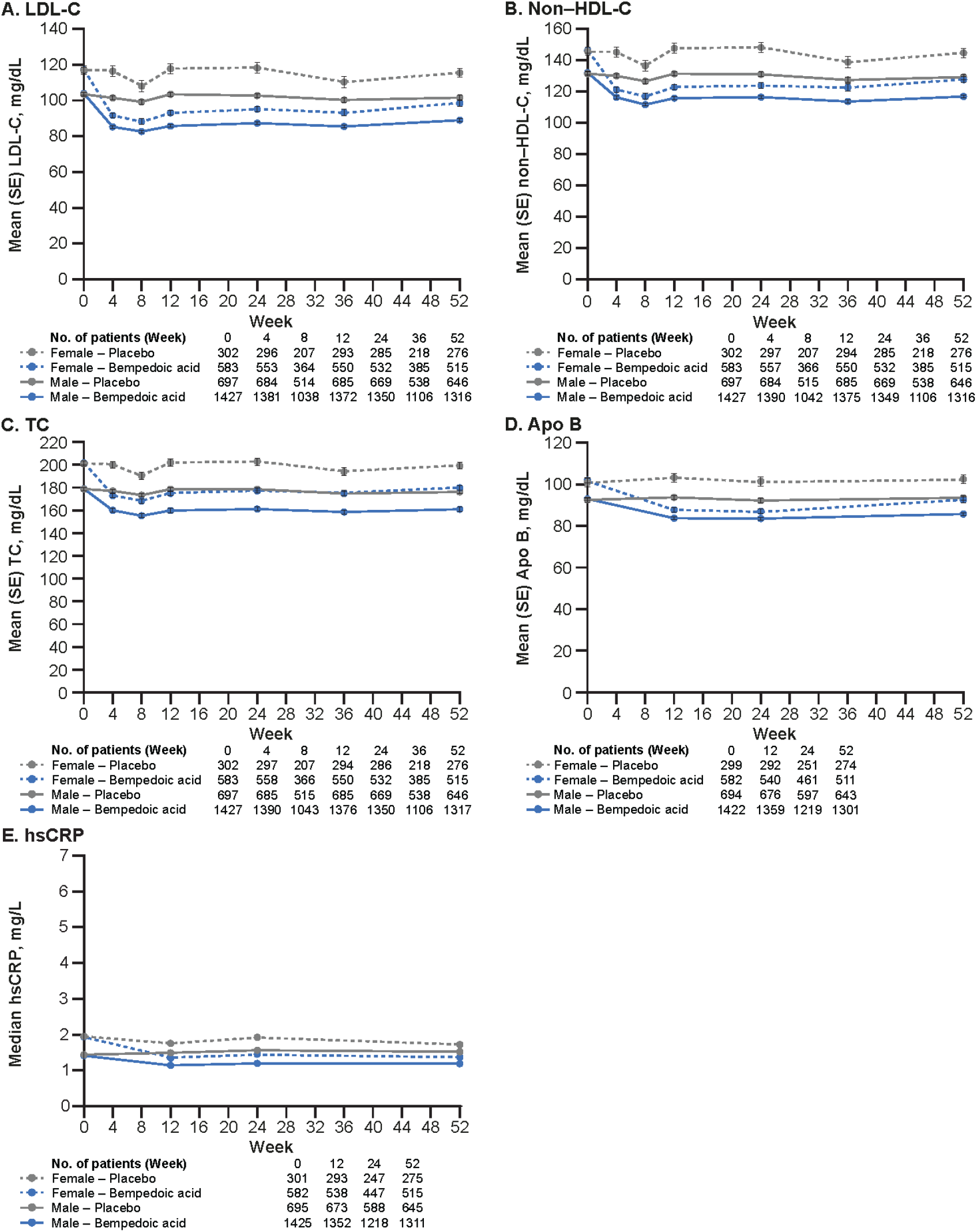
Lipid parameters and hsCRP over time in the ASCVD and/or HeFH on statins pool by treatment and sex at birth. Trends in (A) LDL-C, (B) non–HDL-C, (C) TC, (D) Apo B, and (E) hsCRP from baseline to week 52 in the ASCVD and/or HeFH on statins pool by sex and treatment. Apo B, apolipoprotein B; ASCVD, atherosclerotic cardiovascular disease; HeFH, heterozygous familial hypercholesterolemia; hsCRP, high-sensitivity C-reactive protein; LDL-C, low-density lipoprotein cholesterol; non–HDL-C, non–high-density lipoprotein cholesterol; SE, standard error; TC, total cholesterol.

### Safety

The incidence of TEAEs was generally comparable between the bempedoic acid and placebo arms in both pools (**Table 1**). Among patients receiving bempedoic acid, TEAEs were reported in 77.3% of women and 75.9% of men in the ASCVD and/or HeFH pool and in 60.3% of women and 53.2% of men in the low-dose or no statin pool.

**Table 1.**
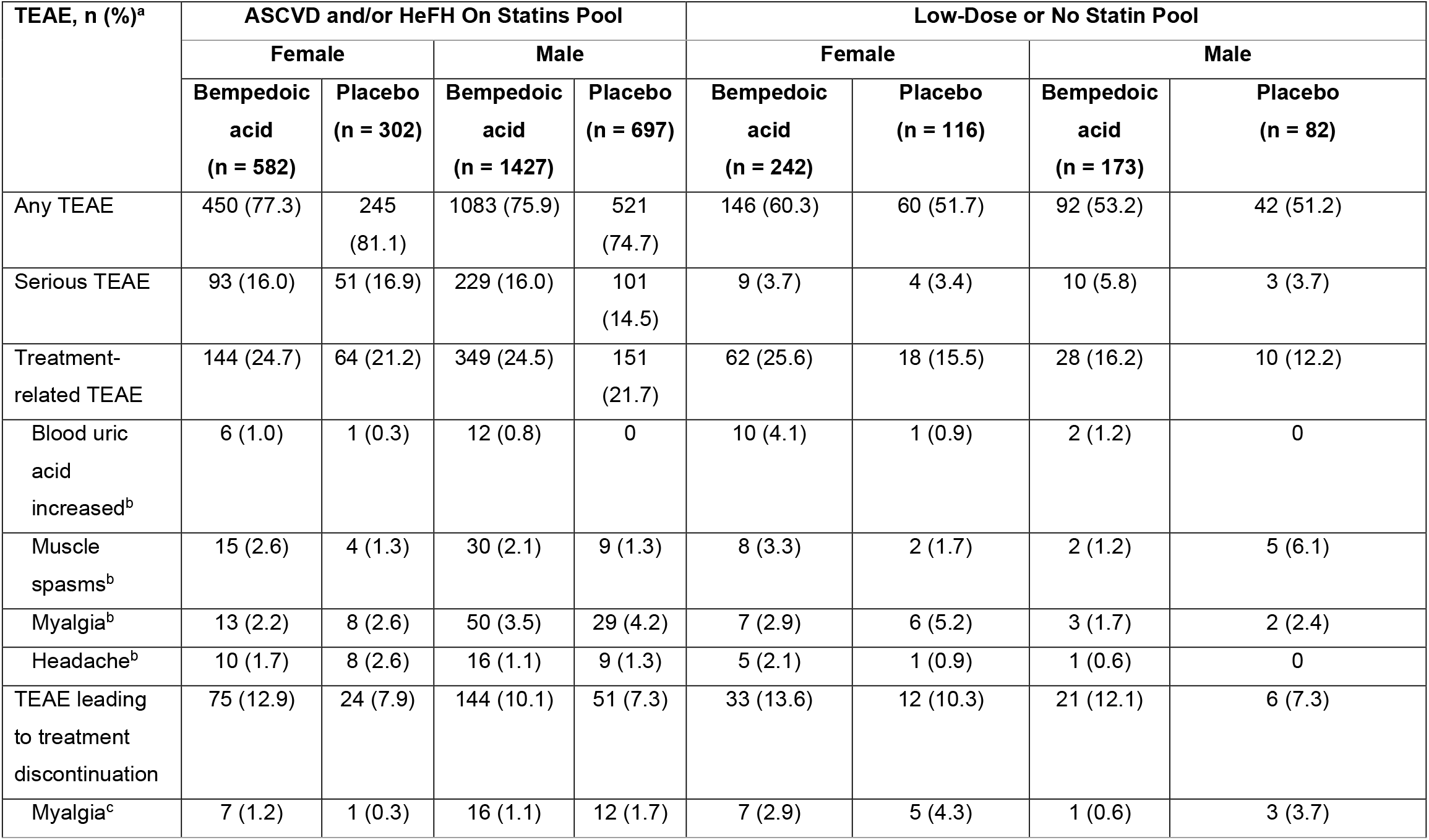
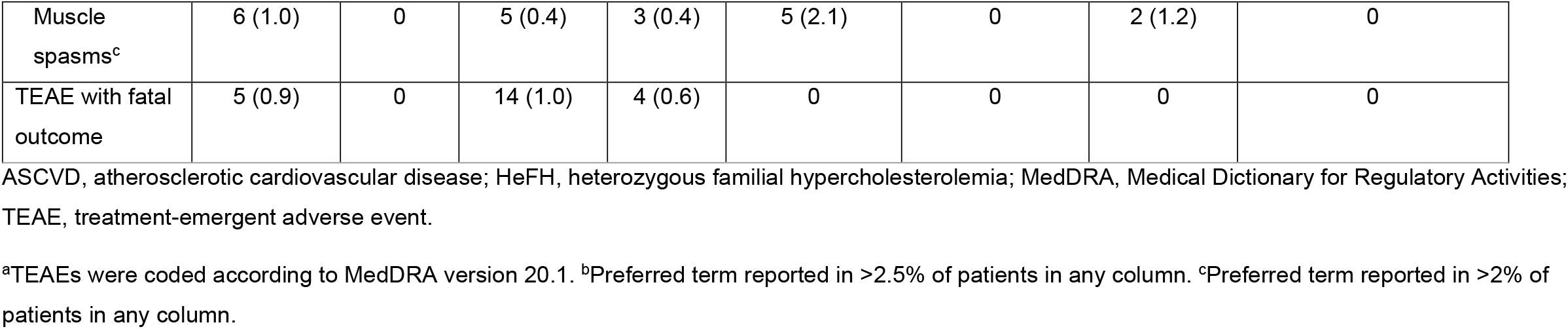
Safety overview by statin pool and sex at birth.

TEAEs leading to discontinuation of bempedoic acid were reported in 12.9% of women and 10.1% of men in the ASCVD and/or HeFH on statins pool and in 13.6% of women and 12.1% of men in the low-dose or no statin pool. The differences between men and women could not be attributed to any specific adverse events. In the low-dose or no statin pool, the incidence of bempedoic acid–related TEAEs was 25.6% in women and 16.2% in men, and was not driven by any one specific type of TEAE. Among patients in the low-dose or no statin pool receiving bempedoic acid, 2.9% and 2.1% of women and 0.6% and 1.2% of men discontinued treatment due to myalgia and muscle spasms, respectively.

In the ASCVD and/or HeFH on statins pool, among the most common TEAEs reported across both treatment groups in women were urinary tract infection (UTI), nasopharyngitis, and upper respiratory infection, and in men were nasopharyngitis, myalgia, and arthralgia (Supplementary Table 3). Across treatment groups in the low-dose or no statin pool, the most common TEAEs in women were UTI, muscle spasms, and myalgia, and in men were muscle spasms, myalgia, and fatigue. In the ASCVD and/or HeFH on statins pool, the incidence of UTI was 9.3% and 9.6% among women receiving bempedoic acid and placebo, respectively; the corresponding incidence in men was 3.0% and 3.3%. In the same pool, headache was reported in 4.1% and 5.0% of women receiving bempedoic acid and placebo; the corresponding incidence in men was 2.2% and 2.3%. Comparable results were observed in the low-dose or no statin pool. The incidence of pain in extremity was higher with bempedoic acid than placebo regardless of sex or statin pool.

The incidences of TEAEs of special interest were generally comparable between women and men in both treatment arms in the two statin pools (Supplementary Table 4). In the ASCVD and/or HeFH on statins pool, new-onset diabetes/hyperglycemia was reported in 3.3% and 7.3% of women receiving bempedoic acid and placebo, respectively; the corresponding incidence in men was 4.6% and 5.3%. In the low-dose or no statin pool, new-onset diabetes/hyperglycemia was reported in 2.9% and 2.6% of women treated with bempedoic acid and placebo, respectively; the corresponding incidence in men was 2.3% and 6.1%. The incidence of muscle disorders was low and generally comparable between men and women receiving bempedoic acid regardless of statin pool. Among patients receiving bempedoic acid in the ASCVD and/or HeFH on statins pool, pain in extremity was reported in 4.5% and 2.5% of women and men, respectively. The corresponding incidence in patients receiving placebo was 3.3% and 1.0%, respectively. The incidence of increased blood creatine phosphokinase among patients in the same pool receiving bempedoic acid was 0.9% among women and 2.4% among men compared with 1.0% and 1.9%, respectively, in the placebo group.

## Discussion

This patient-level, pooled, post-hoc analysis demonstrated a difference in observed reductions of LDL-C levels at week 12 between women and men treated with bempedoic acid. Overall, treatment with bempedoic acid significantly lowered LDL-C and other lipid parameters compared with placebo in both sexes, both in the ASCVD and/or HeFH on statins pool and in the low-dose or no statin pool. Women had significantly greater placebo-corrected reductions than men in LDL-C, non–HDL-C, TC, and Apo B with bempedoic acid in the ASCVD and/or HeFH on statins pool.

Both women and men receiving bempedoic acid in the ASCVD and/or HeFH on statins pool were more likely to achieve LDL-C <100 mg/dL at week 12 than those receiving bempedoic acid in the low-dose or no statin pool. Baseline statin use is unlikely to have contributed to the significantly greater LDL-C reductions observed in women *vs*. men in the ASCVD and/or HeFH pool given that baseline statin intensity was similar between women and men in this pool. Significantly greater LDL-C reductions were also observed in women *vs*. men taking no statin. Moreover, previous subgroup analyses of the same four phase 3 clinical trials reported greater decreases in LDL-C levels with bempedoic acid *vs*. placebo in patients with ASCVD and/or HeFH receiving maximally tolerated statins, regardless of baseline statin intensity, and smaller decreases in LDL-C associated with bempedoic acid *vs*. placebo in statin users compared with non-users at baseline among patients with statin intolerance [20]. In addition, real-world evidence from the PharmLines Initiative database in The Netherlands, showed no significant differences in mean percent changes in LDL-C and TC with statins between women and men [23]. Differences observed in bempedoic acid effects are thus unlikely to stem from background statin therapy.

The greater improvements in LDL-C and other lipid parameters with bempedoic acid observed in women compared with men are in contrast to data from studies of the PCSK9 inhibitors, evolocumab and alirocumab. The Further Cardiovascular Outcomes Research with PCSK9 Inhibition in Subjects with Elevated Risk (FOURIER) cardiovascular outcomes trial reported a smaller reduction in LDL-C with evolocumab in women compared with men at 4 weeks (52% *vs*. 58%, respectively) [24]. Nonetheless, the primary endpoint of cardiovascular death, myocardial infarction, stroke, or hospitalization for unstable angina or coronary revascularization was significantly reduced in both men and women (interaction *p*=0.477), suggesting that the effect of evolocumab on cardiovascular outcomes was not affected by sex [24]. A pooled analysis of 10 ODYSSEY randomized phase 3 trials of alirocumab reported similar results in patients with comparable LDL-C baseline levels to those observed in the current study, with women having smaller LDL-C reductions than men with alirocumab in placebo-controlled trials (−48.3% *vs*. −60.0%), and in ezetimibe-controlled trials (−42.3% *vs*. −50.9%). Furthermore, fewer women than men achieved LDL-C levels <50 mg/dL (36.5% *vs*. 58.7%) with alirocumab [25]. Consistent with results from the FOURIER study [24], for each 39 mg/dL decrease in LDL-C, the difference in risk reduction for major adverse cardiovascular events with alirocumab between women (33%) and men (22%) was not significantly different (*p* for heterogeneity = 0.46) [25]. Similarly, a meta-analysis of 27 trials of statin therapy reported that for every 1 mmol/L (equivalent to ~39 mg/dL [25]) reduction in LDL-C with statins, the proportional and absolute effects of statins on major vascular events were comparable between men and women (*p* for heterogeneity by sex = 0.33) [10].

Bempedoic acid was generally well tolerated. Among patients receiving bempedoic acid in the low-dose or no statin pool, TEAEs were reported in 60.3% and 53.2% of women and men, respectively. In the same pool, 2.1% and 1.2% of women and men, respectively, discontinued bempedoic acid due to muscle spasms, and 2.9% and 0.6% of women and men, respectively, discontinued due to myalgia. The difference between sexes was also noted in the ASCVD and/or HeFH on statins pool but was less pronounced.

A meta-analysis of large, randomized trials of statin therapy showed that women were more likely to experience muscle pain or weakness with moderate-intensity or less intensive statin regimens than men [13]. In a subanalysis of the Understanding Statin Use in America and Gaps in Patient Education (USAGE) survey, the odds of patients reporting muscle symptoms while taking a statin were significantly higher in women than in men (odds ratio, 1.28), as were the odds of discontinuing a statin due to muscle symptoms (odds ratio, 1.48) [12]. In contrast to statins, bempedoic acid is converted to its active metabolite, bempedoyl-CoA, by ASCVL1 in the liver [15] and not in the muscle; hence, muscle symptoms may be less likely. The effect of bempedoic acid is being assessed in the recently completed CLEAR Outcomes, a large cardiovascular outcomes trial (N = 14,014), which will provide an opportunity to gain insights into the long-term cardiovascular clinical outcomes with bempedoic acid and may further elucidate the potential of sex-dependent treatment effects given that 48% of randomized patients are women [26, 27].

There are several limitations in our analyses. This study was a post-hoc analysis of pooled data from four randomized controlled trials, which were not originally designed to determine sex-specific differences. The small number of patients in the low-dose or no statin pool may have hindered the detection of significant differences in treatment effects between women and men. The majority of patients were White, which may limit the generalizability of the results. Furthermore, any differences in either the efficacy or safety of bempedoic acid in premenopausal *vs*. postmenopausal patients could not be reliably determined, given the small proportion of premenopausal patients (≤5.2%). The effect of HRT could not be ascertained due to the low proportion of patients receiving HRT (≤10%). Further investigation of the effect of bempedoic acid in these different groups of women is warranted.

## Conclusions

Treatment with bempedoic acid significantly lowered LDL-C, other lipid parameters, and hsCRP compared with placebo, in both women and men who received maximally tolerated statins. Greater placebo-corrected reductions that were also statistically significant were observed in LDL-C and other lipid parameters in women *vs*. men among patients with ASCVD and/or HeFH taking statins. Bempedoic acid was also generally well tolerated in both women and men and may be a clinically useful non-statin option for patients needing additional LDL-C lowering, including those who are unable to tolerate statins.

## Data Availability

All data produced in the present study are available upon reasonable request to the authors.

## Conflicts of interest

A.C.G. has received research grants/support from Amgen, Akcea/Ionis, Arrowhead, Esperion Therapeutics, Inc., NewAmsterdam Pharma, Novartis, Pfizer, Regeneron, and Sanofi; has served as a consultant for Akcea, Esperion Therapeutics, Inc., NewAmsterdam Pharma, IONIS, Novartis, and Regeneron, and performed editorial work for the Merck Manuals.

M.B. has received research grants/support from Amgen, Sanofi, Mylan/Viatris, and Valeant, and has served as a consultant for Amgen, Daiichi Sankyo, Esperion Therapeutics, Inc., Freia Pharmaceutici, Herbapol, Kogen, KRKA, Novartis, Novo Nordisk, Polfarmex, Polpharma, Regeneron, Sanofi Aventis, Servier, Teva, and Zentiva; he serves as Chief Medical Officer at Nomi Biotech Corporation Ltd.

A.L.C. has received research grants/support from Amarin, Amgen, Menarini, Mylan, Sanofi, and Sanofi Regeneron, and has served as a consultant for or received honoraria from Akcea, Amarin, Amgen, Daiichi Sankyo, Esperion Therapeutics, Inc., Ionis, Kowa, Medco, Menarini, MSD, Mylan, Novartis, Recordati, Regeneron, Sanofi, and The Corpus.

P.B.D. has received institutional research grants/support from Regeneron, Regenxbio, and Retrophin/Travere, and has served as a consultant for Akcea, Esperion Therapeutics, Inc., Ionis, Kaneka, Regeneron, and Novo Nordisk.

L.A.L. has received research grants/support from Amgen, AstraZeneca, Kowa, The Medicines Company, and Novartis. He has also served as an advisor and/or provided continuing medical education on behalf of Amarin, Amgen, AstraZeneca, Esperion Therapeutics, Inc., HLS, Merck, Novartis, Pfizer, and Sanofi.

J.C.H. is an employee of Esperion Therapeutics, Inc. and may own Esperion stock or stock options.

L.L. is a consultant contracted by Esperion Therapeutics, Inc. and may own Esperion stock or stock options.

G.B.J.M. has received research grant(s)/support from Amgen, AstraZeneca, Bayer, Boehringer Ingelheim, HLS Therapeutics, Merck, Novo Nordisk, and Sanofi, and has served as a consultant for these companies, as well as Esperion Therapeutics, Inc., Novartis, Amgen, Sanofi, and Servier.

## Financial support

This work was supported by Esperion Therapeutics, Inc. The work of ALC was supported in part by Ministero della Salute, Ricerca corrente.

## Author contributions

ACG and ALC conceptualized and designed the study. LL performed the statistical analysis. All authors participated in data interpretation, critically reviewed and approved the final manuscript draft submitted for publication, and agree to be accountable for all aspects of the work, ensuring the accuracy and integrity of the publication.

## Acknowledgments

Medical writing and editing assistance were provided by Katerina Pipili, PhD, and Agata Shodeke, PhD, of Spark Medica Inc, funded by Esperion Therapeutics, Inc.

## Notes

### Author Declarations

In all four trials (CLEAR Wisdom [NCT02991118], CLEAR Harmony [NCT02666664], CLEAR Tranquility [NCT03001076], and CLEAR Serenity [NCT02988115]) protocols were approved by local independent ethics committees at each study site, and all study participants provided written informed consent.

