## Supplementary information for "Efficacy and safety of bempedoic acid in women with hypercholesterolemia: Pooled analyses from phase 3 trials"

**SUPPLEMENTARY MATERIAL**

**FIGURES**

**Supplementary Figure 1.** Percent change from baseline to week 12 and placebo-corrected difference (95% CI) in LDL-C in the low-dose or no statin pool by baseline statin intensity and sex at birth. Numbers on the x axis represent the total number of patients within each group with available LDL-C data at week 12.

CI, confidence interval; LDL-C, low-density lipoprotein cholesterol; LS, least squares; SE, standard error.

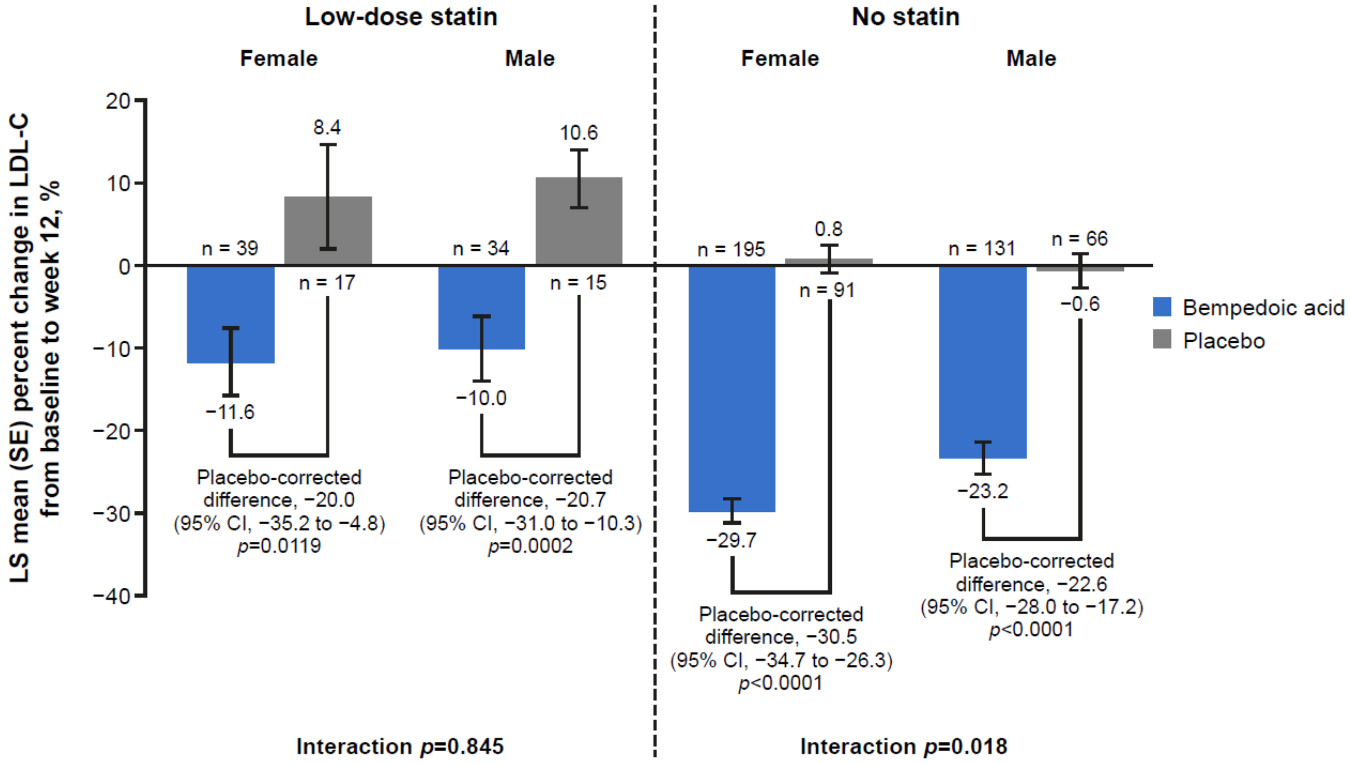

### TABLES

**Supplementary Table 1**. Baseline statin intensity categories (daily dose) [20, 22].

| Pool | High Intensity | Moderate Intensity | Low Intensity[a] |
|---|---|---|---|
| **ASCVD and/or HeFH on statins** | Atorvastatin (40–80 mg) | Atorvastatin (10–20 mg) | Simvastatin (10 mg) |
| | Rosuvastatin (20–40 mg) | Rosuvastatin (5–10 mg) | Pravastatin (10–20 mg) |
| | | Simvastatin (20–40 mg) | Lovastatin (20 mg) |
| | | Pravastatin (40–80 mg) | Fluvastatin (20–40 mg) |
| | | Lovastatin (40 mg) | Pitavastatin (1 mg) |
| | | Fluvastatin XL (80 mg) | |
| | | Fluvastatin (40 mg BID) | |
| | | Pitavastatin (2–4 mg) | |
| **Low-dose or no statin** | **Low Dose** | **Very Low Dose** | |
| | Rosuvastatin (5 mg) | Rosuvastatin (<5 mg) | |
| | Atorvastatin (10 mg) | Atorvastatin (<10 mg) | |
| | Simvastatin (10 mg) | Simvastatin (<10 mg) | |
| | Lovastatin (20 mg) | Lovastatin (<20 mg) | |
| | Pravastatin (40 mg) | Pravastatin (<40 mg) | |
| | Fluvastatin (40 mg) | Fluvastatin (<40 mg) | |
| | Pitavastatin (2 mg) | Pitavastatin (<2 mg) | |

Abbreviations: ASCVD, atherosclerotic cardiovascular disease; BID, twice daily; HeFH, heterozygous familial hypercholesterolemia.

[a]Also includes alternate regimens (ie, every other day or a prespecified number of times/week).

**Supplementary Table 2.** Baseline demographics and clinical characteristics by statin pool and sex at birth.

| Characteristics | ASCVD and/or HeFH on Statins Pool (N = 3008) | | | | Low-dose or No Statin Pool (N = 613) | | | |
|---|---|---|---|---|---|---|---|---|
| | Female | | Male | | Female | | Male | |
| | Bempedoic acid (n = 582) | Placebo (n = 302) | Bempedoic acid (n = 1427) | Placebo (n = 697) | Bempedoic acid (n = 242) | Placebo (n = 116) | Bempedoic acid (n = 173) | Placebo (n = 82) |
| Age, y | 65.7 ± 8.6 | 65.8 ± 8.3 | 65.2 ± 9.2 | 66.4 ± 8.9 | 64.9 ± 10.4 | 63.9 ± 10.8 | 64.2 ± 9.8 | 65.6 ± 8.9 |
| Race, n (%) | | | | | | | | |
| White | 538 (92.4) | 285 (94.4) | 1375 (96.4) | 675 (96.8) | 219 (90.5) | 98 (84.5) | 157 (90.8) | 72 (87.8) |
| Black | 33 (5.7) | 15 (5.0) | 33 (2.3) | 12 (1.7) | 15 (6.2) | 14 (12.1) | 12 (6.9) | 6 (7.3) |
| Asian | 5 (0.9) | 2 (0.7) | 13 (0.9) | 6 (0.9) | 5 (2.1) | 1 (0.9) | 4 (2.3) | 2 (2.4) |
| Other[a] | 6 (1.0) | 0 | 6 (0.4) | 4 (0.6) | 3 (1.2) | 3 (2.6) | 0 | 3 (3.7) |
| Hispanic ethnicity, n (%) | 34 (5.8) | 16 (5.3) | 33 (2.3) | 14 (2.0) | 36 (14.9) | 16 (13.8) | 20 (11.6) | 10 (12.2) |
| BMI, kg/m$^2$ | 30.2 ± 5.8 | 30.3 ± 5.9 | 29.6 ± 4.6 | 29.5 ± 4.5 | 29.2 ± 5.4 | 30.6 ± 5.8 | 30.8 ± 5.1 | 30.5 ± 4.9 |
| History, n (%) | | | | | | | | |
| ASCVD | 542 (93.1) | 284 (94.0) | 1409 (98.7) | 690 (99.0) | NR | NR | NR | NR |
| HeFH | 41 (7.0) | 22 (7.3) | 35 (2.5) | 14 (2.0) | NR | NR | NR | NR |
| Diabetes | 181 (31.1) | 90 (29.8) | 399 (28.0) | 203 (29.1) | 57 (23.6) | 29 (25.0) | 41 (23.7) | 14 (17.1) |
| Hypertension | 487 (83.7) | 251 (83.1) | 1124 (78.8) | 567 (81.3) | 147 (60.7) | 69 (59.5) | 122 (70.5) | 57 (69.5) |
| Menopause status, n (%) | | | NA | NA | | | NA | NA |
| Premenopausal | 30 (5.2) | 5 (1.7) | | | 11 (4.5) | 4 (3.4) | | |
| Hormonal contraceptives | 2 (0.3) | 1 (0.3) | | | 4 (1.7) | 0 | | |
| Not premenopausal | 552 (94.8) | 297 (98.3) | | | 231 (95.5) | 112 (96.6) | | |
| Postmenopausal | 367 (63.1) | 208 (68.9) | | | 128 (52.9) | 67 (57.8) | | |
| Surgically sterile | 185 (31.8) | 89 (29.5) | | | 103 (42.6) | 45 (38.8) | | |
| HRT use[b] | 28 (4.8) | 13 (4.3) | | | 24 (9.9) | 10 (8.6) | | |
| Background LLT, n (%) | | | | | | | | |
| Statin alone | 492 (84.5) | 246 (81.5) | 1194 (83.7) | 591 (84.8) | 8 (3.3) | 7 (6.0) | 8 (4.6) | 3 (3.7) |
| Statin plus other LLT | 67 (11.5) | 43 (14.2) | 201 (14.1) | 90 (12.9) | 33 (13.6) | 13 (11.2) | 27 (15.6) | 12 (14.6) |
| Other LLT alone | 8 (1.4) | 7 (2.3) | 15 (1.1) | 8 (1.1) | 124 (51.2) | 58 (50.0) | 82 (47.4) | 38 (46.3) |

| | | | | | | | | |
|---|---|---|---|---|---|---|---|---|
| None | 15 (2.6) | 6 (2.0) | 17 (1.2) | 8 (1.1) | 77 (31.8) | 38 (32.8) | 56 (32.4) | 29 (35.4) |
| Baseline statin intensity,[c] n (%) | | | | | | | | |
| None | 23 (4.0) | 13 (4.3) | 32 (2.2) | 16 (2.3) | 201 (83.1) | 96 (82.8) | 138 (79.8) | 67 (81.7) |
| Low | 37 (6.4) | 14 (4.6) | 88 (6.2) | 45 (6.5) | 41 (16.9) | 20 (17.2) | 35 (20.2) | 15 (18.3) |
| Moderate | 219 (37.6) | 124 (41.1) | 591 (41.4) | 280 (40.2) | 0 | 0 | 0 | 0 |
| High | 303 (52.1) | 151 (50.0) | 716 (50.2) | 356 (51.1) | 0 | 0 | 0 | 0 |
| Baseline ezetimibe use, n (%) | 33 (5.7) | 31 (10.3) | 117 (8.2) | 45 (6.5) | 123 (50.8) | 61 (52.6) | 92 (53.2) | 41 (50.0) |
| Cholesterol, mg/dL | | | | | | | | |
| Total | 201.6 ± 43.8 | 201.0 ± 45.1 | 178.9 ± 34.0 | 178.6 ± 35.9 | 240.3 ± 44.8 | 232.9 ± 41.5 | 224.4 ± 43.1 | 218.6 ± 45.6 |
| LDL-C | 117.1 ± 38.8 | 116.9 ± 40.2 | 103.9 ± 28.5 | 103.4 ± 29.3 | 148.7 ± 39.1 | 143.8 ± 36.3 | 142.2 ± 39.1 | 137.9 ± 39.6 |
| HDL-C | 55.4 ± 12.6 | 55.4 ± 12.7 | 47.0 ± 11.1 | 47.3 ± 10.8 | 58.1 ± 15.3 | 57.8 ± 20.2 | 47.8 ± 13.5 | 46.9 ± 12.0 |
| Non–HDL-C | 146.2 ± 43.4 | 145.6 ± 45.3 | 131.9 ± 33.6 | 131.3 ± 34.0 | 182.3 ± 44.6 | 175.1 ± 40.4 | 176.7 ± 42.8 | 171.7 ± 48.1 |
| Triglycerides[d] | 132.8 (104.5, 174.5) | 131.2 (100.0, 177.5) | 127.5 (97.5, 171,5) | 124.5 (96.5, 175.0) | 152.0 (115.5, 204.5) | 135.5 (108.0, 191.5) | 157.0 (112, 220.0) | 162 (122.5, 199.5) |
| Apo B, mg/dL | 102.0 ± 30.9 | 100.9 ± 33.2 | 93.1 ± 24.5 | 92.5 ± 25.1 | 134.1 ± 32.0 | 130.4 ± 28.0 | 132.2 ± 29.0 | 130.0 ± 33.5 |
| hsCRP, mg/L[d] | 1.9 (0.9, 4.0) | 2.0 (1.0, 4.3) | 1.4 (0.7, 3.1) | 1.4 (0.8, 3.2) | 3.0 (1.4, 5.3) | 2.7 (1.3, 5.2) | 2.2 (1.0, 4.0) | 1.0 (0.9, 4.5) |

Apo B, apolipoprotein B; ASCVD, atherosclerotic cardiovascular disease; BMI, body mass index; HDL-C, high-density lipoprotein cholesterol; HeFH, heterozygous familial hypercholesterolemia; HRT, hormone replacement therapy; hsCRP, high-sensitivity C-reactive protein; LDL-C, low-density lipoprotein cholesterol; LLT, lipid-lowering therapy; NA, not applicable; NR, not reported; SD, standard deviation.

Data expressed as mean ± SD, unless stated otherwise. [a]Includes American Indian or Alaska Native, Native Hawaiian or other Pacific Islander, other, and multiple. [b]Includes women who are postmenopausal or surgically sterile. [c]Statin intensity classification for the ASCVD and/or HeFH on statins pool was based on the 2013 American College of Cardiology/American Heart Association [22]. See Supplementary Table 1 for all intensity definitions, including those for the low-dose or no statin pool. **[d]**Data are median (quartile 1, quartile 3).

**Supplementary Table 3.** Most common TEAEs by statin pool and sex at birth.

| TEAE, n (%)[a] | ASCVD and/or HeFH On Statins Pool (TEAEs reported in >3% of patients)[b] | | | | Low-dose or No Statin Pool (TEAEs reported in >2% of patients)[c] | | | |
|---|---|---|---|---|---|---|---|---|
| | Female | | Male | | Female | | Male | |
| | Bempedoic acid (n = 582) | Placebo (n = 302) | Bempedoic acid (n = 1427) | Placebo (n = 697) | Bempedoic acid (n = 242) | Placebo (n = 116) | Bempedoic acid (n = 173) | Placebo (n = 82) |
| Urinary tract infection | 54 (9.3) | 29 (9.6) | 43 (3.0) | 23 (3.3) | 12 (5.0) | 14 (12.1) | 1 (0.6) | 0 |
| Nasopharyngitis | 44 (7.6) | 34 (11.3) | 129 (9.0) | 66 (9.5) | 6 (2.5) | 2 (1.7) | 1 (0.6) | 4 (4.9) |
| Upper respiratory tract infection | 35 (6.0) | 16 (5.3) | 56 (3.9) | 24 (3.4) | 2 (0.8) | 2 (1.7) | 1 (0.6) | 2 (2.4) |
| Bronchitis | 29 (5.0) | 12 (4.0) | 31 (2.2) | 13 (1.9) | 7 (2.9) | 3 (2.6) | 0 | 4 (4.9) |
| Muscle spasms | 26 (4.5) | 7 (2.3) | 47 (3.3) | 16 (2.3) | 11 (4.5) | 3 (2.6) | 5 (2.9) | 5 (6.1) |
| Pain in extremity | 26 (4.5) | 10 (3.3) | 35 (2.5) | 7 (1.0) | 9 (3.7) | 2 (1.7) | 5 (2.9) | 2 (2.4) |
| Myalgia | 25 (4.3) | 13 (4.3) | 79 (5.5) | 40 (5.7) | 8 (3.3) | 6 (5.2) | 6 (3.5) | 4 (4.9) |
| Headache | 24 (4.1) | 15 (5.0) | 32 (2.2) | 16 (2.3) | 10 (4.1) | 5 (4.3) | 2 (1.2) | 1 (1.2) |
| Cough | 23 (4.0) | 5 (1.7) | 32 (2.2) | 22 (3.2) | 2 (0.8) | 2 (1.7) | 2 (1.2) | 2 (2.4) |
| Arthralgia | 21 (3.6) | 16 (5.3) | 62 (4.3) | 36 (5.2) | 10 (4.1) | 2 (1.7) | 7 (4.0) | 3 (3.7) |
| Back pain | 21 (3.6) | 7 (2.3) | 46 (3.2) | 15 (2.2) | 2 (0.8) | 4 (3.4) | 6 (3.5) | 1 (1.2) |
| Osteoarthritis | 21 (3.6) | 14 (4.6) | 25 (1.8) | 17 (2.4) | 1 (0.4) | 3 (2.6) | 1 (0.6) | 1 (1.2) |
| Diarrhea | 20 (3.4) | 8 (2.6) | 57 (4.0) | 29 (4.2) | 2 (0.8) | 1 (0.9) | 3 (1.7) | 1 (1.2) |
| Nausea | 20 (3.4) | 10 (3.3) | 24 (1.7) | 13 (1.9) | 6 (2.5) | 2 (1.7) | 3 (1.7) | 1 (1.2) |
| Dizziness | 16 (2.7) | 14 (4.6) | 57 (4.0) | 26 (3.7) | 8 (3.3) | 0 | 2 (1.2) | 1 (1.2) |
| Angina pectoris | 15 (2.6) | 8 (2.6) | 32 (2.2) | 22 (3.2) | 0 | 0 | 2 (1.2) | 0 |
| Hypertension | 12 (2.1) | 8 (2.6) | 38 (2.7) | 24 (3.4) | 6 (2.5) | 3 (2.6) | 5 (2.9) | 0 |

| TEAE, n (%)[a] | ASCVD and/or HeFH On Statins Pool (TEAEs reported in >3% of patients)[b] | | | | Low-dose or No Statin Pool (TEAEs reported in >2% of patients)[c] | | | |
|---|---|---|---|---|---|---|---|---|
| | Female | | Male | | Female | | Male | |
| | Bempedoic acid (n = 582) | Placebo (n = 302) | Bempedoic acid (n = 1427) | Placebo (n = 697) | Bempedoic acid (n = 242) | Placebo (n = 116) | Bempedoic acid (n = 173) | Placebo (n = 82) |
| Fatigue | 8 (1.4) | 8 (2.6) | 36 (2.5) | 26 (3.7) | 5 (2.1) | 4 (3.4) | 5 (2.9) | 4 (4.9) |
| Blood uric acid increased | 10 (1.7) | 2 (0.7) | 23 (1.6) | 2 (0.3) | 11 (4.5) | 2 (1.7) | 7 (4.0) | 0 |
| Blood creatine phosphokinase increased | 5 (0.9) | 3 (1.0) | 34 (2.4) | 13 (1.9) | 2 (0.8) | 0 | 6 (3.5) | 0 |

Abbreviations: ASCVD, atherosclerotic cardiovascular disease; HeFH, heterozygous familial hypercholesterolemia; MedDRA, Medical Dictionary for Regulatory Activities; TEAE, treatment-emergent adverse event.

[a]TEAEs were coded according to MedDRA version 20.1. [b]TEAEs occurring in >3% of patients in any column. [c]TEAEs occurring in >2% of patients in any column.

**Supplementary Table 4.** TEAEs of special interest by statin pool, treatment group, and sex at birth.

| TEAE, n (%)* | ASCVD and/or HeFH on Statins Pool | | | | Low-dose or No Statin Pool | | | |
|---|---|---|---|---|---|---|---|---|
| | Female | | Male | | Female | | Male | |
| | Bempedoic acid (n = 582) | Placebo (n = 302) | Bempedoic acid (n = 1427) | Placebo (n = 697) | Bempedoic acid (n = 242) | Placebo (n = 116) | Bempedoic acid (n = 173) | Placebo (n = 82) |
| New-onset diabetes/hyperglycemia | 19 (3.3) | 22 (7.3) | 66 (4.6) | 37 (5.3) | 7 (2.9) | 3 (2.6) | 4 (2.3) | 5 (6.1) |
| Hepatic enzyme elevations | 16 (2.7) | 4 (1.3) | 35 (2.5) | 11 (1.6) | 9 (3.7) | 0 | 7 (4.0) | 0 |
| Hypoglycemia | 10 (1.7) | 9 (3.0) | 30 (2.1) | 16 (2.3) | 1 (0.4) | 0 | 0 | 0 |
| Metabolic acidosis | 1 (0.2) | 0 | 0 | 0 | 0 | 0 | 0 | 0 |
| Muscular disorders | 79 (13.6) | 27 (8.9) | 186 (13.0) | 75 (10.8) | 27 (11.2) | 11 (9.5) | 20 (11.6) | 12 (14.6) |
| Muscle spasms | 26 (4.5) | 7 (2.3) | 47 (3.3) | 16 (2.3) | 11 (4.5) | 3 (2.6) | 5 (2.9) | 5 (6.1) |
| Pain in extremity | 26 (4.5) | 10 (3.3) | 35 (2.5) | 7 (1.0) | 9 (3.7) | 2 (1.7) | 5 (2.9) | 2 (2.4) |
| Myalgia | 25 (4.3) | 13 (4.3) | 79 (5.5) | 40 (5.7) | 8 (3.3) | 6 (5.2) | 6 (3.5) | 4 (4.9) |
| Increased blood creatine phosphokinase | 5 (0.9) | 3 (1.0) | 34 (2.4) | 13 (1.9) | 2 (0.8) | 0 | 6 (3.5) | 0 |
| Neurocognitive disorders | 3 (0.5) | 4 (1.3) | 11 (0.8) | 4 (0.6) | 2 (0.8) | 0 | 0 | 1 (1.2) |
| Renal disorders | 14 (2.4) | 1 (0.3) | 45 (3.2) | 12 (1.7) | 8 (3.3) | 2 (1.7) | 2 (1.2) | 0 |
| Uric acid elevations/gout | 26 (4.5) | 4 (1.3) | 71 (5.0) | 11 (1.6) | 14 (5.8) | 3 (2.6) | 10 (5.8) | 0 |
| Gout | 3 (0.5) | 0 | 26 (1.8) | 4 (0.6) | 3 (1.2) | 1 (0.9) | 1 (0.6) | 0 |
| Hemoglobin decreased | 18 (3.1) | 9 (3.0) | 47 (3.3) | 13 (1.9) | 2 (0.8) | 0 | 2 (1.2) | 0 |
| Anemia | 16 (2.7) | 6 (2.0) | 41 (2.9) | 13 (1.9) | 1 (0.4) | 0 | 2 (1.2) | 0 |

ASCVD, atherosclerotic cardiovascular disease; HeFH, heterozygous familial hypercholesterolemia; MedDRA, Medical Dictionary for Regulatory Activities; TEAE, treatment-emergent adverse event. *TEAEs were coded according to MedDRA version 20.1.
